# The role of Lipoprotein(a) and oxidized phospholipids in modifying the effects of aspirin on major cardiovascular events and bleeding in the ASPirin in Reducing Events in the Elderly (ASPREE) randomized clinical trial: Statistical analysis plan

**DOI:** 10.64898/2026.05.17.26353443

**Authors:** Rory Wolfe, Harpreet Bhatia, Paul Lacaze, Suzanne G. Orchard, Alice Owen, Galina Polekhina, Chenglong Yu, Robyn L. Woods, Andrew Tonkin, Sotirios Tsimikas

## Abstract

Lipoprotein(a) (Lp(a)) is associated with atherothrombosis through several mechanisms, including putative antifibrinolytic properties. In the ASPREE randomized trial of daily low-dose aspirin for primary prevention in older adults, 72% of trial participants in Australia provided baseline blood samples from which Lp(a) and related oxidized phospholipids and plasminogen have been measured in a specialized laboratory at University of California San Diego. Recent findings from our group suggest that aspirin may benefit older individuals with genotypes associated with elevated lipoprotein(a). We present an analysis plan to address key hypotheses relating to whether the effects of aspirin on cardiovascular disease might vary based on a person’s measured levels of lipoprotein(a), oxidized phospholipid levels present on protein carriers apoB-100 (OxPL-apoB), Lp(a) (OxPL-apo(a)) and plasminogen (OxPL-PLG), and plasminogen. The analysis plan also articulates safety analyses involving major hemorrhage.

## 1 INTRODUCTION

### 1.1 Summary

This Statistical Analysis Plan (SAP) provides a detailed statement of intended secondary analyses of the ASPREE clinical trial in order to address new hypotheses relating to whether the effects of aspirin might vary based on a person’s levels of lipoprotein(a) (Lp(a)) or oxidized phospholipids (OxPL) present on protein carriers apoB-100 (OxPL-apoB), Lp(a) (OxPL-apo(a)) and plasminogen (OxPL-PLG).

The SAP draws extensively from a successful grant application to the National Heart, Lung and Blood Institute (R01 HL170224-01, October 2022), that funded the measurement of Lp(a) and OxPL in ASPREE participants at UCSD, La Jolla, CA, USA, and a published research article reporting aspirin effects on cardiovascular disease (CVD) events and bleeding in the ASPREE trial (McNeil, Wolfe 2018). The SAP provides further detail on the analytical methods and relevant endpoint definitions.

No changes to this SAP will be made after the investigators become unblinded, i.e. once the measured values for Lp(a), OxPL-apoB, OxPL-apo(a), OxPL-PLG and plasminogen, obtained from laboratory analyses being conducted in the US, are transferred to Australia and linked with ASPREE randomization codes held at the ASPREE Data Centre at Monash University, Melbourne, Australia.

The SAP complies with International Conference on Harmonisation (ICH) E9 Guidelines including ICH E9(R1) for handling intercurrent events such as competing risks, loss to follow-up and compliance with study drug over the duration of follow-up (International Conference on Harmonisation 2020).

### 1.2 Background

Lp(a) is associated with atherothrombosis through several mechanisms, including putative antifibrinolytic properties (Bhatia, Becker 2024). Lp(a) is associated with increased CVD risk in primary prevention settings and in patients on statins and PCSK9 inhibitors in secondary prevention settings. Oxidised phospholipids (OxPL) are generated by free-radical mediated lipid peroxidation, or enzymically by lipoxygenases or cyclooxygenase (COX), and are prothrombotic and atherogenic. Lp(a) is a preferential carrier of OxPL, the content of which in plasma can be measured by the assays for OxPL associated with apolipoprotein B100 and apolipoprotein (a) (OxPL-apoB and OxPL-apo(a), respectively). There are no currently approved pharmacological therapies to treat elevated Lp(a), although novel RNA-based medications are in late-stage trials (Thau 2024, Cho 2025). In primary prevention settings, there is some evidence that the use of aspirin (a COX enzyme inhibitor) in adults with elevated Lp(a) (>30 mg/dL or >75 nmol/L) may confer benefit as discussed below. Furthermore, levels of OxPL present on plasminogen (OxPL-PLG) have been associated with faster rates of fibrinolysis, and protection against atherothrombosis. However, whether they are associated with higher rates of bleeding is not known. Mechanistically, evidence suggests it is COX-2 that is involved in OxPL-mediated anti-inflammatory signalling and the resolution of inflammation. But, at low doses, aspirin is highly selective for platelet COX-1, and largely spares COX-2 in other tissues (such as endothelial cells), because it is rapidly cleared from the circulation and new COX enzymes are synthesized in nucleated cells, although COX-2 may be less spared in the setting of long-term daily use of aspirin (Chiang 2004). Finally, while plasma Lp(a) levels are largely genetically driven (Clarke 2009) there are other factors that are known to influence levels, notably the use of statins that may increase Lp(a) levels (Tsimikas 2020, Schwartz 2022, Xie 2025). In summary, physicians do not currently have enough information to treat older primary prevention patients with aspirin, particularly those with a strong family history of CVD.

The overarching aim of this project is to understand the relationship between aspirin and the atherothrombotic risk factors Lp(a) and OxPL using data from a randomized controlled trial of daily low dose aspirin in older adults. The results will move the field beyond existing evidence based on either post-hoc genetic analyses of prior aspirin trials (Chasman 2009, Lacaze 2022, Yu 2024) and observational studies without randomization to aspirin, based on self-reported use (Bhatia, Trainor 2024, Razavi 2024). By doing so, we will provide evidence for clinicians about whether to recommend aspirin in patients with elevated levels of these risk markers, with the potential for advancing strategies for precision medicine. We hypothesize that elevated Lp(a) will identify a subgroup of individuals that may benefit from aspirin in primary prevention settings and propose to test this hypothesis in the ASPREE randomized trial.

The ASPREE trial was a randomized, double-blind, placebo-controlled clinical trial which evaluated the effect of daily use of 100 mg of enteric-coated aspirin in community-dwelling older adults. The trial was approved by the ethics committee at each participating center, and all the participants provided written informed consent before enrollment. The trial funder (the National Institute on Aging) and an independent data and safety monitoring board, whose members had been appointed by the National Institute on Aging, reviewed reports on the accumulating data at regular intervals. Eligible participants were community-dwelling adults from Australia and the United States who were 70 years of age or older (or ≥65 years of age among blacks and Hispanics in the United States). Participants were required to be free from overt coronary heart disease, overt cerebrovascular disease, atrial fibrillation, a clinical diagnosis of dementia, independence-limiting physical disability, a high risk of bleeding, anemia, and a known contraindication to or inability to take aspirin. Key exclusion criteria were the current regular use of an anticoagulant or antiplatelet medication other than aspirin, a systolic blood pressure of 180 mm Hg or more or a diastolic blood pressure of 105 mm Hg or more, a medical indication for or contraindication to regular aspirin therapy, or the presence of a condition that, in the opinion of the primary care physician, was likely to result in death within 5 years.

Annual in-person visits and medical record reviews were supplemented by regular telephone calls to encourage retention in the trial, study medication adherence and facilitate the collection of clinical data. Detailed, high quality demographic, physiological and clinical data were collected at baseline (prior to randomization) and annually through the ASPREE trial. The ASPREE Healthy Ageing Biobank (ASPREE Biobank) collected, processed and stored blood and urine samples at -80degC or under nitrogen vapour at baseline and 3 years later, from Australian ASPREE participants (Parker 2024). For the majority of participants (approximately two-thirds), their baseline sample was collected prior to commencing randomized study medication. For operational and logistical reasons, some participants had their sample collected after commencement of study drug, typically within a month or two. Nearly all samples were collected within a year of randomization with a mean time for collection after randomisation of 50.2 days (Parker 2024). Written informed consent included separate opt-in questions for biomarker and genetic testing. Fractionated blood and urine were aliquoted into multiple low-volume, barcoded cryotubes for frozen storage within 4 hours of collection. Specially designed and outfitted mobile laboratories provided opportunities for sample collection from people in regional and rural areas. 12,219 participants contributed blood/urine at the first timepoint, 10,617 of these older adults provided 3-year follow-up samples. For this project, baseline EDTA plasma aliquots stored in liquid nitrogen were transferred to La Jolla, CA from Melbourne for 11,975 participants.

### 1.3 Main Aims

We propose to test the hypothesis that aspirin modifies the risk of incident atherothrombotic CVD events in primary prevention for older adults to a different extent in people with higher Lp(a), OxPL-apoB, OxPL-apo(a), or OxPL-PLG levels than in people with lower levels.

We propose the following specific aims to test the Lp(a)/OxPL/aspirin hypothesis using data from the ASPREE trial:

#### Aim 1

To test the hypothesis that aspirin modifies the risk of incident major adverse cardiovascular events in primary prevention for older adults to a different extent in people with higher Lp(a) levels than in people with lower Lp(a) levels.

#### Aim 2

To test the hypothesis that aspirin modifies the risk of incident major adverse cardiovascular events in primary prevention for older adults to a different extent in people with higher OxPL-apoB levels than in those with lower OxPL-apoB levels.

#### Aim 3

To test the hypothesis that aspirin modifies the risk of incident major adverse cardiovascular events in primary prevention for older adults to a different extent in people with higher OxPL-apo(a) levels than in those with lower OxPL-apo(a) levels.

#### Aim 4

To test the hypothesis that aspirin modifies the risk of incident major adverse cardiovascular events in primary prevention for older adults to a different extent in people with higher OxPL-PLG levels than in those with lower OxPL-PLG levels.

#### Aim 5

To examine the net benefit of aspirin in people with high versus low levels of Lp(a), OxPL-apoB, OxPL-apo(a) or OxPL-PLG by considering any enhanced benefits on major adverse cardiovascular events against safety considerations in the form of major hemorrhage.

We also have an exploratory aim to examine whether aspirin modifies the risk of incident major adverse cardiovascular events in primary prevention for older adults to a different extent in people with high plasminogen levels than in those with lower plasminogen levels.

### 1.4 Null hypotheses

Among a primary prevention population aged ≥70 years, treatment with daily aspirin 100mg compared with placebo over an average 4.7-year period:

1. does not prevent major adverse cardiovascular events in individuals in a manner that depends on levels of any of plasma Lp(a), OxPL-apoB, OxPL-apo(a), or OxPL-PLG,
2. increases the risk of bleeding equally in individuals with higher and lower levels of each of Lp(a), OxPL-apoB, OxPL-apo(a), and OxPL-PLG.

### 1.5 Alternative hypotheses

Among a primary prevention population aged ≥70 years, treatment with daily aspirin 100mg compared with placebo over an average 4.7-year period:

1a. has a preventive effect on major adverse cardiovascular events, of increasing magnitude <u>with increasing level</u> of Lp(a), OxPL-apoB and OxPL-apo(a), and has an increased preventive effect above threshold levels of each of these biomarkers,

1b. has a preventive effect, on major adverse cardiovascular events, of increasing magnitude <u>with decreasing level</u> of OxPL-PLG, and has an increased preventive effect below threshold levels of OxPL-PLG,

2a. increases the risk of bleeding <u>less</u> in individuals with higher levels of Lp(a), OxPL-apoB and OxPL-apo(a) than in individuals with lower levels,

2b. increases the risk of bleeding <u>more</u> in individuals with higher levels of OxPL-PLG than in individuals with lower levels.

### 1.6 Randomization process

Participants who had a rate of adherence to pill ingestion, as assessed by pill count, of 80% or greater during a 1-month placebo run-in phase were randomly assigned, in a 1:1 ratio, to receive aspirin or placebo. Randomization was stratified according to trial center and age (65 to 79 years or ≥80 years). The randomization sequence was implemented through a secure online system for study research staff to obtain the next available allocation upon activating a request for an eligible participant.

## 2 METHODS

In ASPREE, clinical source documentation — including clinical notes, hospitalization records, and imaging studies (computed tomographic scans or magnetic resonance images) — was sought for events that were thought to be potential endpoints. Committees comprising clinical experts whose members were unaware of the trial-group assignments were responsible for adjudication of all potential clinical endpoint events. Details are given below on the two main endpoints to be used in this study and further details of the definitions and adjudication processes have been published elsewhere, specifically for CVD events (McNeil, Wolfe 2018), stroke (Wolfe 2018), and bleeding (Margolis 2018).

### 2.1 Major adverse cardiovascular events

The main outcome to be used in this study is major adverse cardiovascular events (MACE) defined as a composite of fatal coronary heart disease (excluding death from heart failure), nonfatal myocardial infarction, or fatal or nonfatal ischemic stroke.

In ASPREE, fatal coronary heart disease was defined as death from myocardial infarction, sudden cardiac death, or any other death in which the underlying cause was considered to be coronary heart disease. The definition of nonfatal myocardial infarction was based on joint guidelines of the European Society of Cardiology and the American College of Cardiology (Alpert 2000). Fatal ischemic stroke was defined as any death in which the underlying cause was an obstruction in the intracranial or extracranial cerebral arterial system. The definition of nonfatal ischemic stroke was based on the World Health Organization definition of rapidly developing clinical signs of focal or global disturbance of cerebral function lasting more than 24 hours (unless interrupted by surgery or death), with no apparent cause other than ischemic cerebrovascular disease (No author listed 1989). This MACE endpoint included the conditions related to ischemia and thrombosis that are most likely to be affected favorably by low-dose aspirin.

### 2.2 Major hemorrhage

For analysis of safety in this study the main outcome to be used is major hemorrhage which was a secondary endpoint of the ASPREE trial and which was defined as a composite of hemorrhagic stroke, symptomatic intracranial bleeding, or clinically significant extracranial bleeding. Clinically significant extracranial bleeding was defined as bleeding that led to transfusion, hospitalization, prolongation of hospitalization, surgery, or death.

### 2.3 Exploratory endpoints

In addition to the main endpoints of MACE and major hemorrhage we will also examine other outcomes of relevance including all CVD events (a secondary endpoint of the ASPREE trial consisting of the first event of nonfatal myocardial infarction, fatal and non-fatal stroke, hospitalization for heart failure and coronary heart disease death), CHD death, CVD death (CHD death and fatal stroke), fatal and nonfatal ischemic stroke, all strokes, or myocardial infarction.

In additional exploratory analyses we will examine a broader set of outcomes including all-cause mortality, cancer, dementia, disability-free survival and persistent physical disability, all prespecified endpoints in ASPREE (McNeil, Woods 2018).

### 2.4 Estimands

For all estimands of interest (International Conference on Harmonisation 2020) the following apply in each case:

#### Population-level summary measure

average treatment effect unless otherwise specified.

**Endpoint** is time from randomization until the first occurrence of one of the primary or secondary endpoints.

**Target population** is independent community-dwelling adults aged 70 years and older free of any history of cardiovascular disease events, attending primary care with their general practitioner.

**Treatment condition** is daily 100mg enteric-coated aspirin tablets for an average 4.7-year period and compared to identical placebo. This includes allowance for temporary interruption to manage tolerability, and allowance for commencement of open-label aspirin treatment according to treating clinician advice in response to a participant experiencing a recognized clinical indication (such as myocardial infarction or stroke).

#### Intercurrent events and strategies to address these

(i) the occurrence of death (for non-fatal endpoints) and certain causes of death (for endpoints that include specific causes of death) will be treated as competing risks; (ii) the withdrawal of consent occurred in only a small number of participants and such individuals will be analyzed on the assumption of withdrawal at random relative to the outcome events being analyzed; (iii) the cessation of aspirin use (for the randomized aspirin group) or commencement of open-label aspirin (for the randomized placebo or aspirin groups) will be dealt with by restricting attention to the estimation of intention-to-treat effects.

### 2.5 Measurement procedure for Lp(a) and OxPL

The previously unthawed aliquots of 0.5 ml plasma were shipped to UCSD in a blinded fashion with the only identifying information being barcodes etched onto the base of the cryotubes. Samples were stored at UCSD at -70°C in a monitored freezer. The UCSD laboratory has extensive expertise in measuring Lp(a) with well-validated, clinically-predictive, isoform-independent Lp(a) assay traceable to the WHO/IFCC SRM-2B reference material that use antibodies developed in the laboratory. For this study established methods were used for Lp(a) (Leibundgut 2012, Marcovina 2022, Marcovina 2026) and OxPL-apoB, OxPL-apo(a), OxPL-PLG and plasminogen (Tsimikas 2020, Clarke 2022, Marcovina 2026). A validated standard curve, as well as high- and low-quality control samples were used to maintain assay rigor.

## 3 DATA ANALYSIS

The data will be analyzed by statisticians based at the Biostatistics Unit, School of Public Health and Preventive Medicine, Monash University. The analyses will be performed using the statistical software packages Stata and R. Two-sided p-values will be used throughout and 0.05 will be taken as the cut-off for statistical significance.

### 3.1 Description of cohort

Characteristics of the two randomized treatment groups at randomization will be tabulated, overall and further stratified by dichotomized levels of each of Lp(a) and OxPL measures. For these tables we will use mean and standard deviations for continuous measures that follow approximately a symmetrical distribution on their scale of measurement or after logarithm transformation, or medians and interquartile ranges otherwise, and numbers and percentages for categorical measures. No p-values will be calculated for this table since a null hypothesis of no difference between the two groups at baseline (i.e. that the two groups are random samples from the same underlying population) is known to be true due to the randomization process that was used in the trial.

Given the large sample size, randomization is anticipated to adequately balance baseline characteristics of participants in the two treatment groups. Hence unadjusted analyses will be considered primary.

Measured levels of plasma Lp(a), OxPL-apoB, OxPL-apo(a), OxPL-PLG and plasminogen will be compared between individuals according to presence of LPA SNPs and by their LPA polygenic risk score.

### 3.2 Comparisons between randomized groups

Comparisons will be made between treatment arms on an intention-to-treat basis, that is, according to the group to which participants were randomized and regardless of adherence to assigned treatment.

#### 3.2.1 Lp(a) and aspirin’s effect on MACE

The relationship between Lp(a), randomized treatment group, their interaction and time to the first occurrence of each endpoint of interest (MACE, all CVD events, etc.) will be assessed using Cox proportional hazards regression analysis. Each endpoint will be analyzed in a separate time-to-event analysis using a Cox model. Because the distribution of Lp(a) is right skewed, a log transformation will be applied and log-base2 will be used to enhance interpretability of relevant model coefficients. If there are any zero values then these will be replaced using half the lower limit of detection, prior to log transformation. Event rates (time to first event within each endpoint definition) will be calculated and compared using hazard ratios and 95% confidence interval from the Cox models. To test the study’s main hypotheses for Lp(a) (Sections 1.4 and 1.5) each model will include two covariates and their interaction: the first covariate will be an indicator of the group to which the individual was randomized, aspirin or placebo, the second covariate will relate to plasma Lp(a) level. The p-value for the interaction between randomized group and the Lp(a) variable will provide the means to directly test the study hypothesis in each model.

Three separate main analyses will be performed with the Lp(a) variable respectively defined as: (i) continuous log-Lp(a), (ii) a binary indicator of Lp(a) >75 nmol/L (∼30 mg/dL), and (iii) a binary indicator of Lp(a) > 90^th^ percentile of the study cohort. Stratified analyses of aspirin’s effect will be conducted in the two groups defined by Lp(a) ≤ vs > 75 nmol/L and in the two groups defined by 0-90^th^ percentile vs > 90^th^ percentile, each presented with the corresponding interaction p-value.

For MACE, the time to event is defined as time from randomization date to the date of occurrence of that event (i.e. the endpoint date), as confirmed by the adjudication committee. For participants who do not experience the endpoint of interest, the administrative censoring date will be the earliest of date of death, loss to follow up, or the end date of the trial’s intervention phase.

In analyses of MACE, death due to causes other than those specified by the endpoint will be considered as censoring events, i.e. cause-specific hazard ratios will be estimated, and for graphical representation these will be treated as competing risks and cumulative incidence plots will be shown with cumulative incidence estimated using the Aalen–Johansen estimator (Aalen 1978).

Secondary analyses will examine Lp(a) < vs. ≥125 nmol/L (∼50 mg/dL), and Lp(a) as deciles to examine the possibility of non-linearities such as a J-shaped relationship and to identify the possibility of an optimal threshold for identifying a subgroup of participants who may benefit from aspirin. The log-Lp(a) linear association assumption made in the main analyses will be tested by comparison to a Lp(a) linear association and against spline-based (non-linear) models.

The proposed Lp(a) cutoffs are validated for risk prediction in primary prevention and used in in clinical laboratories, guidelines and expert consensus statements (Erqou 2009, Nordestgaard 2010, Willeit 2018, Mach 2020, Pearson 2021).

#### 3.2.2 OxPL-apoB and aspirin’s effect on MACE

To test hypotheses relating to OxPL-apoB (Sections 1.4 and 1.5), the same analysis methods will be followed as in Section 3.2.1 with Lp(a) being replaced by OxPL-apoB. OxPL-apoB will be log(base2)-transformed. Two separate main analyses will be performed with the OxPL-apoB variable respectively defined as: (i) continuous log-OxPL-apoB, (ii) a binary indicator of > 80^th^ percentile of OxPL-apoB in the study cohort. Stratified analyses of aspirin’s effect will be conducted in the two groups defined by OxPL-apoB 0-80^th^ percentile vs > 80^th^ percentile. Secondary analyses will use OxPL-apoB threshold of 8.5 nmol/L, representing the 75^th^ percentile in CVD populations (Gilliland 2023), and decile analyses.

#### 3.2.3 OxPL-apo(a) and aspirin’s effect on MACE

To test hypotheses relating to OxPL-apo(a) (Sections 1.4 and 1.5), the same analysis methods will be followed as in Section 3.2.1 with Lp(a) being replaced by OxPL-apo(a). OxPL-apo(a) will be log(base2)-transformed. Two separate main analyses will be performed with the OxPL-apo(a) variable respectively defined as: (i) continuous log-OxPL-apo(a), (ii) a binary indicator of > 80^th^ percentile of OxPL-apo(a) in the study cohort. Stratified analyses of aspirin’s effect will be conducted in the two groups defined by OxPL-apo(a) 0-80^th^ percentile vs > 80^th^ percentile. Secondary analyses will use OxPL-apo(a) threshold of 37.5 nmol/L, representing the 75^th^ percentile in CVD populations (Gilliland 2023), and decile analyses.

#### 3.2.4 OxPL-PLG and aspirin’s effect on MACE

To test hypotheses relating to OxPL-PLG (Sections 1.4 and 1.5), the same analysis methods will be followed as in Section 3.2.1 with Lp(a) being replaced by OxPL-PLG. The main analyses to be performed are with: (i) continuous OxPL-PLG, and (ii) a binary indicator of > 80^th^ percentile of OxPL-PLG in the study cohort. Stratified analyses of aspirin’s effect will be conducted in the two groups defined by OxPL-PLG 0-80^th^ percentile vs > 80^th^ percentile. Secondary analyses will investigate OxPL-PLG in deciles. In additional analyses for OxPL-PLG we will add plasminogen to the factors to be adjusted for (see Section 3.3.2).

#### 3.2.5 Plasminogen and aspirin’s effect on MACE

In exploratory analyses, the same methods will be followed as in Section 3.2.1 with Lp(a) replaced by plasminogen. The main analyses to be performed are with: (i) continuous plasminogen, and (ii) a binary indicator of > 80^th^ percentile of plasminogen in the study cohort. Stratified analyses of aspirin’s effect will be conducted in the two groups defined by plasminogen 0-80^th^ percentile vs > 80^th^ percentile. Secondary analyses will investigate plasminogen in deciles. In additional analyses for plasminogen we will add OxPL-PLG to the factors to be adjusted for (see Section 3.3.2).

#### 3.2.6 Aspirin’s effects on major hemorrhage and other endpoints

The entire set of analyses described in sections 3.2.1 to 3.2.5 will be repeated for major hemorrhage (in place of MACE) and then for each of the other endpoints specified in Section 2.3.

The pre-specified interaction p-values from the main analyses will be presented only for MACE and major hemorrhage, and there will be no adjustment for multiplicity. All other endpoints that are analyzed (see list in Section 2.3) will be considered exploratory. For all analyses, cause-specific hazard ratios (for main effects) and ratios of cause-specific hazard ratios (for interaction effects) will be presented with 95% confidence intervals and without p-values.

### 3.3 Additional analyses and checking assumptions

#### 3.3.1 Assumption checking

The proportional hazards assumptions of the main analyses will be tested using Schoenfeld residuals, and a p-value for the test will be presented. The tests will have excessively high power in most cases and thus to avoid over-identification of minor departures from non-proportional hazards a plot of log(-log) transformed estimated survival probabilities against log-transformed at-risk time will be assessed visually for evidence of parallel lines for the two treatment groups. If the proportional hazards assumption is considered inappropriate, then a sensitivity analysis will be undertaken to examine the evidence for a time-dependent effect of aspirin.

#### 3.3.2 Adjusted analyses

A secondary set of analyses will be performed using multivariable Cox proportional hazards regression models to adjust for the following set of baseline characteristics that are thought to be strongly prognostic of outcome in this population: age at randomization, sex, smoking, hypertension, diabetes, non-HDL-C, and statin use (a set of confounders commonly used in relevant epidemiological studies), and an extended set that additionally includes HDL-C, alcohol consumption, body mass index, previous regular aspirin use, chronic kidney disease (eGFR < 60 ml/min/1.73m^2^ or urinary albumin:creatinine ratio > 3 mg/mmol), liver function and nonsteroidal anti-inflammatory drug use at baseline. We will employ multiple imputation for missing data if more than 5% of participants have missing data in one or more of these prognostic variables; we will use complete-case analysis if <5% of participants have missing data.

#### 3.3.3 Net benefit analyses

The main analyses will be extended to undertake a net benefit analysis to examine whether there is a subgroup of older individuals with elevated Lp(a) in whom the benefits of low-dose aspirin (in reducing MACE) outweigh the harms of bleeding. This analysis will also be further extended to incorporate longer-term effects on MACE and bleeding through use of data from the ASPREE-XT post-trial extended follow-up study.

#### 3.3.4 Restricted mean survival time analyses

Difference in restricted mean survival time (RMST) or restricted mean time lost (RMTL – if censoring due to death or competing causes of death is relevant) between randomized groups will be calculated as a treatment effect to be reported, either alongside the corresponding hazard ratio or, in cases where the proportional hazards assumption is found to be violated, in preference to an overall hazard ratio. RMST analyses will use a Royston-Parmar flexible parametric model with three degrees of freedom for the baseline hazard and two degrees of freedom for a time-dependent covariate effect.

#### 3.3.5 Analyses utilizing biomarker combinations

Further analyses will be undertaken to test for “Atherothrombotic risk”: Lp(a) + OxPL-apo(a), and “Hemostasis/bleeding risk”: plasminogen + OxPL-PLG. In these analyses, we will examine combinations of two of the biomarkers simultaneously with a hierarchy of models evaluated for improvements of model fit using likelihood-ratio tests: (i) a model with no biomarkers, (ii) a model with one biomarker, (iii) a model with two biomarkers additively, and (iv) a model with two biomarkers multiplicatively (through an interaction term). Categorical cross-classification of these pairs of biomarkers will also be examined using the threshold cutoffs from the main analyses and following the approach we have used previously (Bhatia, Dweck 2024). Ratios of two biomarkers will also be examined, specifically OxPL-apo(a)/Lp(a) for atherothrombotic risk and OxPL-PLG/plasminogen for bleeding risk. We will also undertake exploratory analyses using the ratios of OxPL-apoB/OxPL-PLG, OxPL-apo(a)/OxPL-PLG, OxPL-apoB/OxPL-apo(a) and OxPL-apoB/Lp(a).

#### 3.3.6 Other analyses

Analyses that are undertaken and reported but which are not included in this SAP will be labelled in all reports as not being pre-specified (i.e. as being post-hoc).

### 3.4 Sensitivity analyses

Sensitivity analyses will be undertaken in the form of subgroup analyses by (i) diabetes status at baseline and (ii) an indicator of whether a participant’s baseline blood sample was taken before or after commencement of study medication.

For the main endpoints, a time-dependent analysis will be undertaken to incorporate the potential influence of changing use of prescribed statins during follow-up. This analysis will adjust for factors thought likely to influence commencement of statins in this age group including but not limited to time-dependent reports of cardiovascular-related events that were not substantiated as endpoints, familial cardiovascular disease history and comorbidity burden.

Further sensitivity analyses of MACE will estimate sub distribution hazard ratios using the Fine and Gray approach to allow for the competing risk of non-MACE mortality.

### 3.5 Power calculation

In an estimated 11,985 participants with available plasma samples there were approximately 413 MACE events, 544 “all” CVD events, and 569 deaths during the ASPREE clinical trial. To compare MACE risk between two groups of participants divided at Lp(a) of 75 nmol/L (assuming this identifies the top quintile), there is 80% power to detect approximately twice as strong an effect of aspirin on MACE risk in the Lp(a) ≥75 nmol/L group than the <75 nmol/L group. With Lp(a) as a continuous variable there is power to detect smaller interaction effects.

### 3.6 Analysis populations

The intention to treat analysis set will include all participants randomized and whose blood samples were successfully measured for Lp(a).

### 3.7 Ethics approval

Consent for use of participant biospecimen samples was obtained from participants at the time of biospecimen collection (Project 18/08 - Alfred Hospital Human Research Ethics Committee).

Ethics approval was obtained (Project 245/21, approval date 22 April 2021) from the Alfred Hospital Human Research Ethics Committee for analysis of ASPREE participant biospecimens to quantify a panel of atherogenic and thrombosis-related biomarkers including lipoprotein(a), OxPL-apoB, OxPL-apo(a), OxPL-PLG and associated biomarkers, and to examine their association with cardiovascular and cancer outcomes and possible associations with effects of aspirin.

## Data Availability

All data used in the present work are available after publication of the main results in a peer-review journal and upon reasonable request to the ASPREE Governance Board in the form of an expression of interest submitted through the ASPREE Access Management Site (AMS): https://ams.aspree.org/public/

